# Exploration of Modern Contraceptive Methods Using Patterns among Late Reproductive Aged Women in Bangladesh

**DOI:** 10.1101/2023.08.23.23294471

**Authors:** Md Shohel Rana, Md Badsha Alam, Md Tahir Hassen, Md Iqbal Kabir, Shimlin Jahan Khanam, Md Nuruzzaman Khan

## Abstract

**Background:** In light of the increasing population of late reproductive-aged women (aged 35 and above) within the broader demographic of reproductive-aged females, the concern surrounding their contraceptive considerations has escalated to a point of critical importance. This study aims to examine the trends and determinants of modern contraceptive practices among late reproductive-aged women in Bangladesh.

**Methods:** A total of 17,736 women aged 35 and more were included in the analysis, utilizing data from three consecutives Bangladesh Demographic and Health Surveys conducted in 2011, 2014, and 2017-18. The outcome variable was the use of modern contraceptive methods (yes or no). The explanatory variables encompassed survey years, individual characteristics of the women, as well as characteristics of their partners and the community. To examine the association of the outcome variable with the explanatory variables, a multilevel logistic regression model was used.

**Results:** Approximately 54% of women aged 35 and older do not utilize modern contraceptive methods, and there have been no significant shifts in their usage observed over the survey years. The probability of using modern contraceptive methods exhibited a notable decline in relation to increasing age, the educational level of women’s partners, and their categorization within the richer or richest wealth quintile. Conversely, women with higher education, increased exposure to mass media, and residing in the Dhaka or Rajshahi division displayed an elevated likelihood of embracing modern contraceptive methods.

**Conclusion:** The study highlights the relatively stable adoption of modern contraceptive methods among women aged 35 or older in Bangladesh, despite their increasing representation within the population. This raises concerns about the elevated risk of unintended pregnancies and shorter birth intervals, emphasizing the need for targeted interventions to address the specific needs and preferences of this demographic.

## Introduction

The ongoing public health challenges in low- and middle-income countries (LMICs) are primarily attributed to the higher occurrence of unintended pregnancy, short inter-pregnancy intervals, and a higher rate of abortions [1–3]. Access to modern contraceptive methods can effectively address these issues by empowering women to exercise control over their reproductive choices, enabling them to space pregnancies and plan the timing of childbirth [4, 5]. This, in turn, contributes to healthier birth outcomes and reduces the risk of complications associated with closely spaced pregnancies [6, 7]. Moreover, by addressing them, modern contraception tackles one of the key drivers of maternal mortality, as women who have access to and utilize contraception are more likely to access maternal healthcare services due to counseling by family planning workers [8, 9]. Additionally, the utilization of modern contraceptives is linked to a decrease in high-risk pregnancies, such as those among adolescents and older women, resulting in lower rates of preterm births and low birth weights [4]. The use of modern contraceptives methods also facilitates family planning, enabling couples to make informed decisions about the number of children they wish to have, thus aiding in the optimization of maternal and child health resources [10]. These interventions collectively contribute to achieving Sustainable Development Goals (SDG) 3, which aims to ensure healthy lives and promote well-being for all, and SDG 5, which targets gender equality and women’s empowerment [11, 12]. The contributions of modern contraceptive methods to reducing maternal mortality (70 per 100,000 live births) and child mortality (12 and 25 per 1,000 live births for neonatal and under-five mortality, respectively) in line with the SDG targets are also recognized worldwide [13, 14].

The focus on contraceptive practices has traditionally centered on women in their prime reproductive years below 35 years. However, as demographics shift and societal dynamics evolve, an emerging demographic subset is garnering increased attention worldwide – late reproductive-aged women (aged 35 years or more) [15, 16]. In LMICs, this age-specific phenomenon assumes an even more compelling dimension due to demographic transitions, resulting in a larger number of women residing within this age range [17, 18]. Furthermore, considering the rising trend of delayed childbearing and changing family dynamics in LMICs, the implications of contraceptive utilization among late reproductive-aged women become increasingly pertinent [10, 19]. Women in this age group might be at an increased risk of medical comorbidities and complications during pregnancy, making the adoption of effective contraception a pivotal consideration for their well-being. However, elderly reproductive-aged women in LMICs may encounter distinctive challenges when seeking contraception [20]. Cultural expectations surrounding fertility and family structure might influence their decisions, potentially leading to overlooked reproductive health concerns [1, 14]. Additionally, limited healthcare infrastructure and resources in LMICs, along with their intentional exclusion of comparatively older women, can hinder access to appropriate contraceptive methods and comprehensive reproductive healthcare services for this demographic [3, 9]. Therefore, gaining a comprehensive understanding of the factors influencing contraceptive practices among elderly reproductive-aged women and how these practices have changed over the years is of paramount importance [21].

However, despite its critical importance, this issue remains largely unexplored in LMICs, including Bangladesh. Existing studies have primarily focused on either young women or the entire reproductive-aged female population and examine socio-demographic factors associated with contraception use [4, 5, 22–24]. As a result, the dynamics of contraception among late reproductive aged women remains unknown. This is a significant concern, particularly in Bangladesh, where the black hole effect of population momentum results in a larger number of women entering the 35+ age group each year, surpassing the number of women entering reproductive age [8, 25]. This insufficient attention might be linked with the stagnation of contraception use rate that Bangladesh has witnessed over the years [10, 26]. To address this gap, the current study aims to explore the contraceptive dynamics among late reproductive aged women in Bangladesh and assess how these dynamics have evolved across different survey periods. Furthermore, the study seeks to identify factors associated with modern contraceptive methods use among these late reproductive aged women.

## Methods

### Data source and sampling

The data were derived from three consecutives Bangladesh Demographic and Health Surveys (BDHSs) conducted in 2011, 2014, and 2017-18. Detailed information about the methodologies employed in the BDHSs can be located in the corresponding survey reports [27–29]. In summary, these surveys encompassed nationally representative samples of women within the reproductive age bracket (15–49 years), selected through a two-stage stratified random sampling process. In the initial stage, 600 enumeration areas (clusters) for the 2011 and 2014 surveys, and 672 enumeration areas for the 2017-18 survey, were randomly chosen as primary sampling units, based on the National Population and Housing Census conducted in 2011 by the Bangladesh Bureau of Statistics. During the subsequent stage, an average of 30 households per enumeration area were selected via systematic random sampling, culminating in a total of 17,964 households in 2011, 17,989 households in 2014, and 20,160 households in 2017-18. The interviews were carried out in 17,141 households (n=17,842 women) in 2011, 17,300 households (n=17,863 women) in 2014, and 19,457 households (n=20,127 women) in 2017-18.

### Study sample

A total of 17,736 women were encompassed in this study, comprising 5,479 women from 2011, 5,505 from 2014, and 6,752 women from the 2017-18 BDHS, as per the predetermined inclusion criteria. These inclusion criteria included being married or in a union, aged between 35 and 49 years, being fecund, currently not being pregnant or within the postpartum amenorrhea period and do not want a baby within two years of the survey date.

### Outcome variable

The primary outcome for this study was the use of modern contraceptive methods. The relevant data was derived by asking two subsequent questions. Initially, eligible women were asked, “*Are you or your husband currently using any method to delay or avoid getting pregnant?*” The responses were reported dichotomously as either yes or no. If women provided an affirmative response, they were subsequently asked, “*Which method are you using?*” To answer this question, women were presented with a list of contraceptive methods’ names: pills, injections, implants, IUDs, condoms, female sterilization, male sterilization, periodic abstinence, and withdrawal. Additionally, an open option was provided if the contraception used was not listed. We reclassified these responses into two categories: “*modern contraceptive methods users*” and “*others,*” in accordance with the World Health Organization’s classification [30]. Modern contraceptive methods include pills, injections, implants, Intra Uterine Devices (IUDs), condoms, and female and male sterilization. Individuals who did not use any contraception or used traditional methods were collectively classified as “*others.*”

### Explanatory variables

Aligned with the study’s objective, the year of the survey constituted a primary explanatory variable. Other explanatory variables were selected based on a comprehensive literature search, as well as their availability in the analyzed surveys, and their statistical significance in relation to modern contraceptive use [4, 31–35]. The selected variables were women’s age (35-39, 40-44, 45-49 years), women’s education levels (no education, primary, secondary, higher), women’s employment status (unpaid work, paid work), and number of ever-born children (≤2 children, >2 children). Partner’s educational attainment (no education, primary, secondary, higher), partner’s occupation categories (agriculture, physical worker, services, business, others), household type (nuclear, joint), and wealth index were also included. The survey authority created wealth index variable through principles component analysis of the variables covering several households assets, including ownership radio, television and household’s roof type. Other variables included were place of residence (urban, rural), and region (Barisal, Chattogram, Dhaka, Khulna, Rajshahi, Rangpur, Sylhet).

### Statistical analysis

Descriptive analysis was used to depict the characteristics of study participants and the distribution of modern method contraceptive use across the explanatory variables considered. The association of each explanatory variable with the outcome variable was examined using the chi-square test. Multilevel logistic regression model was used to explore the associations of outcome variable with explanatory variables. The utilization of multilevel modeling stemmed from the hierarchical structure of the BDHSs data, where individuals are nested within households and households are nested within communities. In this study design, an additional layer is introduced, representing the year of the survey. Previous studies have shown that for such data structures, multilevel modeling yields superior outcomes compared to conventional logistic regression models [36]. Multicollinearity was assessed as part of the analysis. The outcomes are presented as adjusted Odds ratios (aOR) accompanied by their corresponding 95% confidence intervals (95% CI). All statistical analyses were conducted using Stata software version 14 (Stata Corp, College Station, Texas, USA).

### Ethics Approval

The data analyzed in this study were obtained from the Demographic and Health Survey Program of the USA. Prior to conducting the survey in Bangladesh, approval was obtained from the institutional review board of ICF, USA, and subsequently from the National Research Ethics Committee of the Bangladesh Medical Research Council. To ensure the participants’ consent, informed written consent was acquired from all individuals involved, utilizing an appropriate institutional form. These consent forms were securely archived by the survey authority. In our research, we sought permission to access the data for analytical purposes, and the survey authority provided us with non-identified data. As the study involved secondary data analysis and adhered to the relevant guidelines and regulations, no additional ethical approval was required.

## Result

### Background characteristics of the respondents

Table 1 presents background characteristics of the respondents, while year wise distribution is presented in Supplementary table 1. The percentage of women within the late reproductive age cohort was found to be 37.4% during the 2011 BDHS, a figure that slightly increased to 38.6% and 39.6% during the BDHS conducted in 2014 and 2017-18, respectively. The cumulative proportion of these late reproductive-aged women was 38.7%. In 2011, 54.1% of late reproductive-aged women possessed an educational attainment at or above the primary level. This percentage exhibited an incremental progression, reaching 57.6% and 68.8% in the 2014 and 2017-18 BDHSs. Approximately three-quarters of the total women analysed reported rural areas as their place of residence. Another noteworthy trend was the ascent in the percentage of women who reported engaging in paid employment, indicating a rapid increase across the triad of surveys – from 11.0% in 2011 to 38% in 2014, culminating at 56% in the 2017-18 iteration.

**Table 1.** Selected background characteristics of the late reproductive aged women, N=17,736.

| Characteristics | Overall |  |
| --- | --- | --- |
|  | Frequency(n) | Percentage (%) |
| **Women's age (in years)** |  |  |
| 35-39 | 6,851 | 38.7 |
| 40-44 | 5,895 | 33.2 |
| 45-49 | 4,990 | 28.1 |
| **Women's education** |  |  |
| No education | 6,989 | 39.4 |
| Primary | 5,908 | 33.3 |
| Secondary | 3,694 | 20.8 |
| Higher | 1,145 | 6.5 |
| **Women's working status** |  |  |
| Unpaid work | 11,260 | 63.5 |
| Paid work | 6,476 | 36.5 |
| **Women's partner education** |  |  |
| No education | 6,429 | 36.3 |
| Primary | 4,810 | 27.2 |
| Secondary | 4,137 | 23.2 |
| Higher | 2,360 | 13.3 |
| **Women's partner occupation** |  |  |
| Agriculture | 6,203 | 35.0 |
| Physical worker | 5,400 | 30.4 |
| Services | 1,305 | 7.4 |
| Business | 3,839 | 21.6 |
| Others | 989 | 5.6 |
| **Type of household** |  |  |
| Nuclear ($\leq 4$) | 7,491 | 42.3 |
| Joint ($> 4$) | 10,245 | 57.7 |
| **Number of children ever born** |  |  |
| ≤2 children | 4,512 | 25.4 |
| >2 children | 13,224 | 74.6 |
| **Wealth Index** |  |  |
| Poorest | 2,985 | 16.8 |
| Poorer | 3,609 | 20.3 |
| Middle | 3,662 | 20.7 |
| Richer | 3,572 | 20.1 |
| Richest | 3,908 | 22.1 |
| **Media exposure** |  |  |
| Unexposed | 7,329 | 41.3 |
| Exposed | 10,407 | 58.7 |
| **Place of residence** |  |  |
| Urban | 4,841 | 27.3 |
| Rural | 12,895 | 72.7 |
| **Region** |  |  |
| Barishal | 1,120 | 6.3 |
| Chittagong | 2,997 | 16.9 |
| Dhaka | 5,563 | 31.4 |
| Khulna | 2,250 | 12.7 |
| Rajshahi | 2,051 | 11.6 |
| Rangpur | 2,300 | 12.9 |
| Sylhet | 1,455 | 8.2 |

**Note:** Established in 2015, the Mymensingh division was recognized as an independent region in the 2017/18 BDHS, following its previous inclusion in the Dhaka division during the 2011 and 2014 BDHSs. To ensure uniform classification, we consequently reclassified the Mymensingh division under the Dhaka division for the 2017/18 BDHS.

### Distribution of contraceptive methods use

A noteworthy 61.83% (10,966) of women were identified as active users of various contraceptive methods, while the remaining 38.17% (6,770) chose not to partake in contraceptive practices as evidenced across the three surveys. A breakdown reveals that 46.52% embraced modern contraceptive methods, whereas 15.31% favored traditional alternatives. Taking into account the comprehensive data, the overarching prevalence of modern contraceptive utilization stood at 46.52%. This figure demonstrated a consistent pattern with values of 46.09% in 2011, 45.97% in 2014, and 47.31% in the 2017-18 survey. Pills accounted for 19.13% of usage, followed by sterilization at 11.04%, and injections at 9.1%.

**Table 2.** Percentage distribution of elderly reproductive aged women by current contraceptive use across the three surveys.

| **Contraceptive Use** | **2011**<br>**(N=5,479)**<br>**%** | **2014**<br>**(N=5,505)**<br>**%** | **2017-18**<br>**(N=6,752)**<br>**%** | **Total**<br>**(N=17,736)**<br>**%** |
| --- | --- | --- | --- | --- |
| **Modern contraceptive method** | **46.09** | **45.97** | **47.33** | **46.52** |
| Pill | 19.43 | 18.97 | 19.02 | 19.13 |
| Injections | 8.43 | 9.58 | 9.29 | 9.11 |
| Condoms | 4.61 | 4.99 | 5.60 | 5.11 |
| Female sterilization<sup>♀</sup> | 10.03 | 8.32 | 9.05 | 9.13 |
| Male sterilization<sup>♀</sup> | 1.80 | 2.06 | 1.89 | 1.91 |
| IUD<sup>♀</sup> | 0.89 | 0.64 | 0.75 | 0.76 |
| Implant<sup>♀</sup> | 0.90 | 1.41 | 1.70 | 1.36 |
| **Traditional contraceptive method** | **15.36** | **13.73** | **16.55** | **15.31** |
| Periodic abstinence | 12.36 | 11.05 | 13.26 | 12.29 |
| Withdrawal | 2.24 | 2.10 | 2.87 | 2.44 |
| Others | 0.75 | 0.59 | 0.44 | 0.59 |
| **Do not use contraception** | **38.55** | **40.30** | **36.12** | **38.17** |
| **Total** | **100.00** | **100.00** | **100.00** | **100.00** |

<sup>♀</sup> High effective contraceptive method

### Distribution of modern contraceptive methods use across respondents’ socio-demographic characteristics

The overall distribution of the utilization of modern contraceptive methods across respondents’ socio-demographic characteristics is presented in Table 3, while year-wise distribution of modern contraceptive methods use is presented Supplementary table 2. Women’s age, educational background, occupational status, husband’s education and occupation, household type, wealth index, and region of residence were identified as pertinent variables that intricately influenced the utilization of modern contraceptive methods.

**Table 3:** Distribution of elderly reproductive aged women reported their current modern contraceptive method use patterns associated with different health and demographic surveys in Bangladesh.

| **Characteristics** | **Modern contraceptive methods use status**<br>**(n=17,736)** |  | **p-value** |
| --- | --- | --- | --- |
|  | **Use** | **Non-use** |  |
| **Women's age (in years)** |  |  | p<0.001 |
| 35-39 | 50.22 | 28.55 |  |
| 40-44 | 32.79 | 33.63 |  |
| 45-49 | 16.99 | 37.82 |  |
| **Women's education** |  |  | p<0.001 |
| No education | 37.84 | 40.77 |  |
| Primary | 33.70 | 32.97 |  |
| Secondary | 21.37 | 20.36 |  |
| Higher | 7.09 | 5.90 |  |
| **Women's working status** |  |  | p<0.001 |
| Unpaid work | 61.09 | 65.57 |  |
| Paid work | 38.91 | 34.43 |  |
| **Women's partner education** |  |  | p<0.001 |
| No education | 38.13 | 34.61 |  |
| Primary | 26.86 | 27.35 |  |
| Secondary | 21.82 | 24.63 |  |
| Higher | 13.19 | 13.41 |  |
| **Women's partner occupation** |  |  | p<0.001 |
| Agriculture | 35.93 | 34.14 |  |
| Physical worker | 29.72 | 31.08 |  |
| Services | 7.37 | 7.35 |  |
| Business | 23.61 | 19.94 |  |
| Others | 3.37 | 7.49 |  |
| **Type of Household** |  |  | p<0.001 |
| Nuclear ($\leq 4$) | 39.92 | 44.25 | |
| Joint ($> 4$) | 60.08 | 55.75 | |
| **No. of ever-born children** |  |  | p<0.01 |
| $\leq 2$ children | 24.13 | 26.58 | |
| $> 2$ children | 75.87 | 73.42 | |
| **Wealth Index** |  |  | p<0.001 |
| Poorest | 17.57 | 16.18 |  |
| Poorer | 21.04 | 19.73 |  |
| Middle | 21.27 | 20.10 |  |
| Richer | 19.49 | 20.71 |  |
| Richest | 20.61 | 23.27 |  |
| **Mass media exposure** |  |  | p<0.05 |
| Unexposed | 40.29 | 42.23 |  |
| Exposed | 59.71 | 57.77 |  |
| **Place of residence** |  |  | p=0.421 |
| Urban | 26.96 | 27.59 |  |
| Rural | 73.04 | 72.41 |  |
| **Region of residence** |  |  | p<0.001 |
| Barishal | 5.95 | 6.63 |  |
| Chattogram | 15.66 | 17.97 |  |
| Dhaka | 41.96 | 39.93 |  |
| Khulna | 12.48 | 12.87 |  |
| Rajshahi | 14.26 | 11.85 |  |
| Rangpur | 7.99 | 8.38 |  |
| Sylhet | 1.70 | 2.37 |  |

### Factors associated with modern contraceptive methods use among late reproductive aged women in Bangladesh

The factors associated with the utilization of modern contraceptive methods were determined using the multilevel logistic regression model, and the results are presented in Table 4. The results of each survey are presented in the Supplementary table 3. We did not report a significant change in the likelihood of modern contraceptive methods use across the survey years. We found declined likelihoods of modern contraceptive method use among women aged 40-45 (aOR=0.53, 95% CI: 0.49, 0.57) and 45-49 (aOR=0.24, 95% CI: 0.22, 0.26) years old as compared to women aged 35-39 years old. In comparison to women with no educational attainment, higher likelihoods (ranging from 112% to 142%) of modern contraceptive methods use were found among women with primary to higher education. An alternative association was observed for partner’s education, where an increasing level of partner’s education was negatively associated with the utilization of modern contraceptive methods. The likelihoods of modern contraceptive methods use were found 17% (0.83, 95% CI, 0.74-0.94) and 24% (aOR=0.76, 95% CI: 0.66, 0.88) lower among women with richer and richest wealth quintile as compared to the women with poorest household wealth quintile. Higher likelihoods of modern contraceptive method use were observed among women who residing in the Dhaka (aOR=1.22, 95% CI: 1.07, 1.38) and Rajshahi (aOR=1.37, 95% CI: 1.19, 1.59) regions as compared to the women in the Barishal region. The likelihood of using modern contraceptive methods was found to be increased by 20% (aOR=1.22; 95%CI: 1.13, 1.32) among those exposed to mass media, compared to those who were not exposed to mass media. Women who reported having paid work were 1.19 times more likely (aOR=1.19; 95%CI: 1.10, 1.28) to use modern contraceptive methods as compared to the women who were not engaged in any paid work. The likelihood of modern contraceptive method use was found 43% higher (aOR, 1.43, 95% CI, 1.32-1.55) among women who had more than 2 children as compared to the women who had at least two children.

**Table 4.**
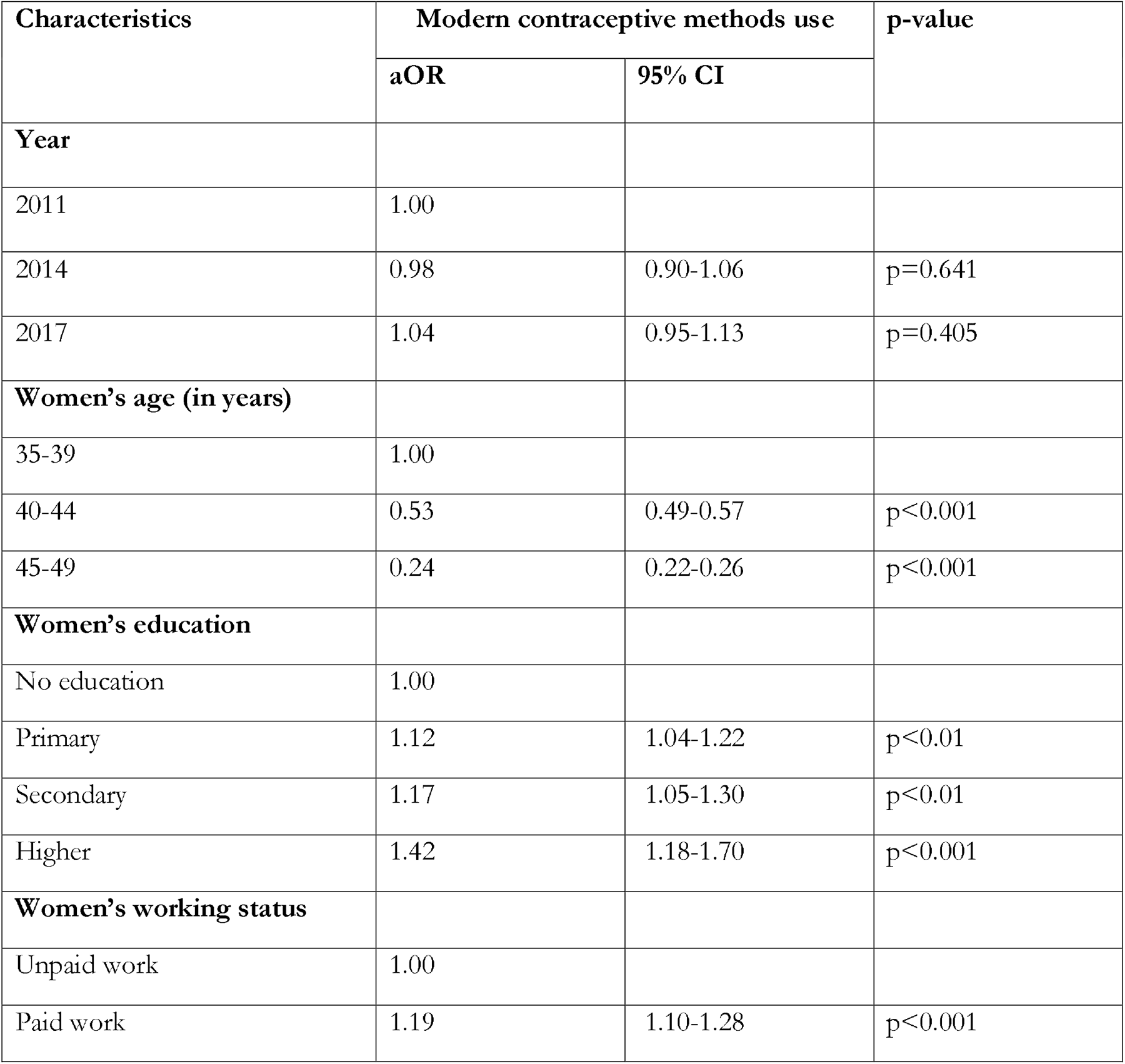
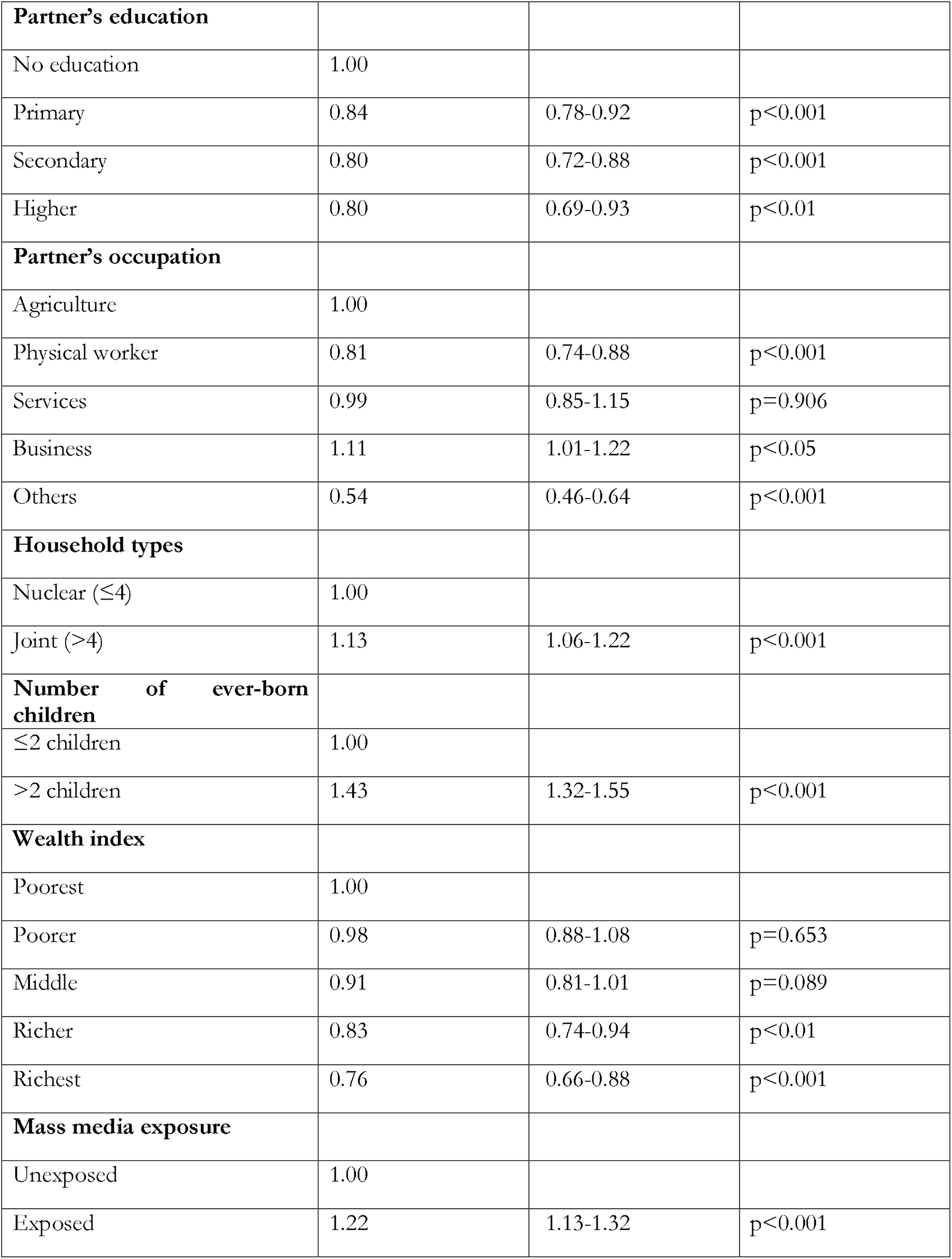
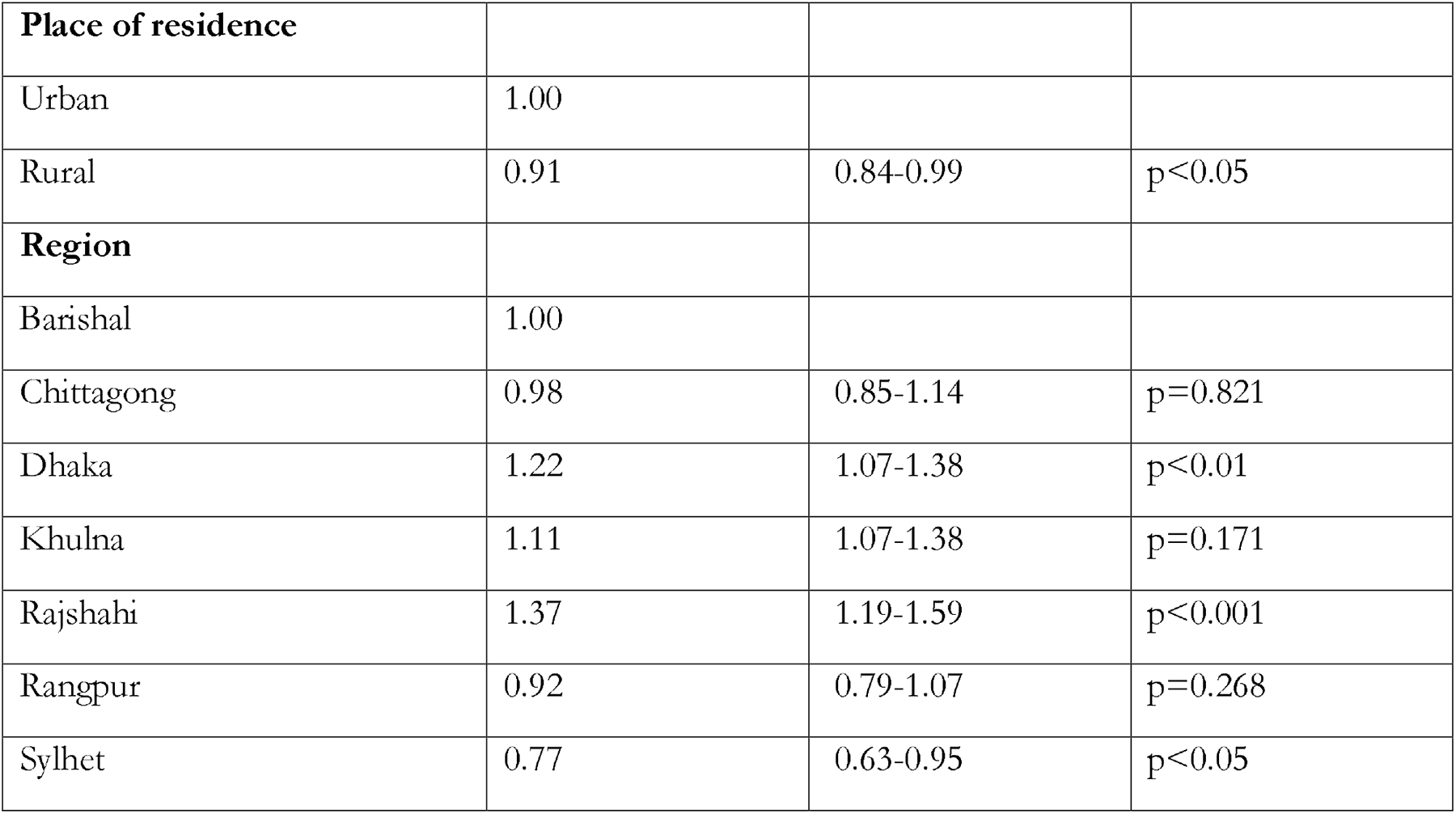
Multilevel logistic regression model to explore likelihoods of modern contraceptive methods use across survey years adjusted for possible covariates, Bangladesh.

## Discussion

The aim of this study was to examine the patterns of contraceptive practices among elderly women of reproductive age in Bangladesh. We also assessed how these trends changed over different survey periods and identified the factors that influence the use of modern contraceptive methods. No notable shifts in the uptake of modern contraceptive methods were observed over the surveyed years. The likelihood of adopting modern contraceptive methods exhibited a decline in correlation with the advancing age of women, as well as with the education level of women’s partners, and their inclusion within the richer or richest wealth quintile. Conversely, a declined likelihood of modern contraceptive methods use was found to be associated with higher levels of women’s education, increased exposure to mass media, and residence in either the Dhaka or Rajshahi division. These findings emphasize that the uptake of modern contraceptive methods remained relatively stable among women aged 35 or older, despite their increasing numbers over the years in Bangladesh. In conjunction with the ongoing rise in the prevalence of pregnancies among late reproductive aged women, this situation suggests that these women are at risk of experiencing unintended pregnancies and shorter birth intervals, both of which could lead to significant adverse consequences. It is crucial to link these observations with the current stagnation in the utilization of modern contraceptive methods in Bangladesh.

The findings of this study revealing a nearly 46% utilization of modern contraceptive methods among elderly women aged 35 or more, arise concern for several reasons, despite the apparent improvement compared to other LMICs [5, 7, 24, 37–40]. The primary apprehension stems from the fact that this particular demographic currently constitutes more than 25% of the total reproductive-aged women in Bangladesh. Furthermore, this percentage is expected to increase in the forthcoming years due to the country’s population structure, where a significant number of women currently fall within the 15 to 30-year age range, and they will eventually transition into the 35+ cohort as time progresses [25]. Importantly, the inadequate increase in modern contraceptive methods use within this growing demographic should be considered one of the major factors contributing to the stagnation of contraception use rates in Bangladesh over the years [41].

Given the current pregnancy dynamics, it is notable that approximately one third of all pregnancies in Bangladesh are either unintended or occur within a short interval [1, 2, 7]. Additionally, over half of these pregnancies involve women aged 35 or older [7]. As the population of women aged 35 and above continues to grow, these numbers are poised to increase further in the coming years. This challenge comes at a time when Bangladesh is undergoing a rapid nutritional transition, coupled with a notably higher prevalence of overweight/obesity among women aged 35 and above [42]. Chronic conditions, such as diabetes and hypertension, are also highly prevalent within this demographic, affecting nearly 40% of all women [23, 43]. A significant proportion of these cases remain undiagnosed, untreated, or uncontrolled. This number is currently experiencing rapid escalation. The convergence of unintended and short interval pregnancies with these chronic health conditions is anticipated to result in severe adverse consequences [6]. These consequences add to the existing burden of unintended and short interval pregnancies, which already contribute to lower usage of maternal healthcare services and a higher prevalence of adverse maternal and child health outcomes, including maternal and child mortality [3, 9]. Moreover, Bangladesh is currently witnessing a rapid shift in fertility age, with an increasing prevalence of pregnancies occurring after the age of 30. This trend is particularly conspicuous among higher-educated and urban women, within whom the prevalence of overweight/obesity and chronic conditions is also notably high. Collectively, these findings underscore a serious challenge that Bangladesh will face in the coming years if proactive measures to ensure contraception among this growing demographic are not implemented [44].

The consistent rate of modern contraceptive method uses among women aged 35 is likely due to a prevailing focus solely on women in earlier reproductive stages (age below 35) [19]. This tendency primarily arises from the misconception within communities that contraception is primarily necessary during the initial phases of reproductive life [44]. This oversight at the service providers’ level is often accompanied by challenges faced by comparatively elderly populations, including a sense of discomfort when seeking contraception [45]. Furthermore, women in this age group often lead busy lives, juggling family responsibilities and occupations, which can lead to contraception being perceived as a lower-priority issue [8, 35]. Our study findings indicating lower likelihoods of modern contraceptive uptake among women in the richest and richer households support this trend. Adding to this, a notable proportion of these women also manage chronic conditions, potentially leading to the misconception that such health issues influence their fertility to the extent that contraception becomes unnecessary [46]. This specific misconception is widespread in Bangladesh, particularly among those who are illiterate and homemakers, and it contributes to the prevailing dynamics within this demographic [8]. Supporting this notion, our study reported higher likelihoods of modern contraceptive method usage among women with comparatively higher education, formal occupations and higher exposure to mass media that increase knowledge about importance of using contraception, similar to other available studies [4, 39].

This study has also reported lower likelihoods of modern contraception uptake as the level of women’s partner education rises. This represents a U-shaped change in the findings, which contrasts with the observation that a higher level of husband education contributes to increased contraception uptake among women of earlier reproductive age [39]. This contrast further indicates a neglect of the importance of contraception use among husbands as women’s age increase. This negligence impacts the overall contraception uptake in this age group, considering that husbands play a significant role as decision-makers for the use or non-use of contraception in Bangladesh [22]. Notably, while women are the primary users of contraception, husbands have a major influence on the decision-making process [38, 44]. The findings related to regional variations in the likelihood of contraception uptake align with the overall pattern of contraception use in Bangladesh. These variations should be correlated with healthcare facility density, the quality of available family planning services, and prevailing social norms [10].

This study has several strengths and a few limitations. Analysis of this study covering multiple survey rounds provides a comprehensive view of how modern contraceptive utilization has evolved over the years among 35+ aged women in Bangladesh. The use of nationally representative data enhances the generalizability of the findings to the broader population of elderly reproductive-aged women in the country, while also ensuring a diverse geographic representation that strengthens the robustness of the results. The study employs a multilevel logistic regression model to explore the associations between various socio-demographic factors and modern contraceptive use among elderly women. This approach helps to account for hierarchical data structures within the BDHS datasets, thereby improving the accuracy of the estimates regarding the factors that influence modern contraceptive use. By including a range of socio-demographic characteristics, the study offers a comprehensive understanding of the multifaceted determinants that shape contraceptive choices among this demographic [25]. However, certain limitations should be acknowledged when interpreting the study findings [47]. BDHSs data are self-reported, as such they may introduce recall bias. Additionally, the cross-sectional nature of the BDHS data restricts the study’s ability to establish causal relationships; while associations can be identified, causation cannot be inferred. The analysis is constrained by the available variables in the BDHS datasets, potentially omitting relevant contextual factors that influence contraceptive decision-making [15]. Furthermore, the study’s findings could not capture the evolving landscape of healthcare access, family planning programs, and societal attitudes towards contraception because of the lack of relevant variables in the data sets [44]. Despite these limitations, the study provides valuable insights into the trends and determinants of modern contraceptive use among elderly reproductive-aged women in Bangladesh, offering a foundation for future research and policy considerations.

## Conclusion

Nearly 54% of women in Bangladesh aged 35 and older do not use modern contraceptive methods, with no significant shifts observed over the surveyed years. The likelihood of using modern contraceptive methods declines notably with increasing age, partner’s education level, and wealth quintile. Conversely, an increased likelihood of embracing modern contraceptive methods was observed among women with higher education, increased exposure to mass media, and residence in Dhaka or Rajshahi division. These findings highlight the stable adoption of modern contraceptive methods among women aged 35 or older, despite their growing representation in the population. The persistent trend of stagnation calls for proactive measures to address the specific needs of elderly reproductive aged women in family planning programs. Strengthening awareness campaigns, improving healthcare access, and tailoring interventions to their preferences could lead to a more effective and responsive approach to contraception among elderly reproductive aged women.

## Abbreviation

BDHS: Bangladesh Demographic and Health Survey
SDG: Sustainable Development Goals
MDG: Millenium development Goals
aOR: Adjusted Odds Ratio
TFR: Total Fertility Rate
IUD: Intrauterine Device
LMIC: Low-and Middle-Income Country
CI: Confidence Interval

## Acknowledgement

The authors thank the MEASURE DHS for granting access to the 2011 and 2017/18 BDHS data.

## Authors contribution

MSR and MNK developed the study concept. The formal data analysis conducted by MSR and write the primary draft of the manuscript along with MBA. MTH, MNK, SJK and MIK reviewed the manuscript carefully. All authors reviewed and approved the final version of the manuscript.

## Data availability

The datasets used and analysed in this study are available from the Measure DHS website: https://dhsprogram.com/data/available-datasets.cfm

## Acknowledgments

The contributors would to convey their deep gratitude for the assistance they have granted by the Department of Population Science at Jatiya Kabi Kazi Nazrul Islam University, Bangladesh, where this investigation was carried out.

